# Cohort Profile: Prematurity Immunology in HIV-infected Mothers and their infants Study (PIMS)

**DOI:** 10.1101/2020.03.18.20033654

**Authors:** Thokozile R Malaba, Clive M Gray, Landon Myer, Marie-Louise Newell, for the PIMS Study Group

## Abstract

**Purpose:** PIMS, is a prospective cohort study in South Africa investigating the association between antiretroviral therapy (ART) use, preterm delivery (PTD)and small-for gestational age (SGA) live births. PIMS main hypotheses are that ART initiation in pregnancy and ART-induced hypertension are associated with PTD and SGA respectively and that reconstitution of cellular immune responses in women on ART from before pregnancy results in increases in PTD of appropriate-for-gestational age (AGA) infants.

**Participants:** Pregnant women (n=3972) aged ≥18 years regardless of HIV status recruited from 2015 to 2016 into the overall PIMS cohort (2517 HIV-uninfected, 1455 HIV-infected). A nested cohort contained 551 HIV-infected women who were ≤24 weeks’ GA on ultrasound: 261 initiated ART before pregnancy, 290 initiated during pregnancy.

**Findings to date:** Women in the overall cohort were followed antenatally through to delivery using routine clinical records; further women in the nested cohort were actively followed up until 12 months postpartum, with data were collected on maternal health (HIV care and ART use, clinical care and inter-current clinical history). Other procedures conducted on the nested cohort included physical examinations (anthropometry, blood pressure measurement), assessment of fetal growth (ultrasound), maternal and infant phlebotomy for storage of plasma, RNA and peripheral blood mononuclear cells, collection of delivery specimens (placenta and cord blood), and infant 12 month developmental assessment. Preliminary findings have contributed to our understanding of risk factors for adverse birth outcomes, and the relationship between pregnancy immunology, HIV/ART and adverse birth outcomes.

**Future plans:** Using specimens collected from HIV-infected study participants throughout pregnancy and first year of life, the PIMS provides a valuable platform for answering a variety of research questions focused on temporal changes of immunology markers in women whose immune status is altered by HIV infection, and how ART initiated during pregnancy affects immune responses. The relationship between these immunological changes with adverse birth outcomes as well as possible longer-term impact of exposure to ART in fetal and early life will be explored. Additionally, further active and passive follow-up of mothers and their infants is planned at school-going age and beyond to chart growth, morbidity and development, as well as changes in family circumstances.

## Introduction

Antiretroviral drugs in HIV-infected pregnant women prevent mother-to-child transmission (PMTCT) and delay HIV disease progression. WHO guidelines now recommend antiretroviral therapy (ART) for all, immediately upon HIV diagnosis, including for HIV-infected women during pregnancy and breastfeeding, to be continued lifelong ^1^. However infants born to HIV-infected mothers would be exposed to multi-drug ART regimen for prolonged periods at a crucial time during their development 2, which could result in decreased health, developmental, and survival outcomes ^3 4^. In high maternal HIV prevalence settings, the increasing population of ART-exposed infants could make the goal of under-five mortality reduction less likely.

Adverse birth outcomes contribute significantly to under-five mortality, as well as infant health and developmental problems ^5^. There is an ongoing debate regarding the association between exposure to maternal ART during fetal life and adverse birth outcomes, following reports from Europe ^6-9^, USA 10 and Africa ^11-14^ of possible ART-associated increased risk of preterm delivery (PTD), small-for-gestational age (SGA) or low birthweight (LBW) infants. Furthermore, these exposed infants are also at increased risk of acquiring viral infections ^15 16^, as well as the negative impact of ART on fetal brain development and function ^17^. Interpretation of findings from various studies, especially from African settings, is hindered by the reliability of gestational assessment, with ultrasound dating in early pregnancy usually unavailable.

There is limited understanding of general pregnancy-related immune changes in high HIV prevalence settings. Successful pregnancies require intricate fetal-maternal (FM) immune balances, to enable maternal tolerance of the semi-allogeneic fetus; this FM tolerance is primarily maintained by the placenta ^18 19^. Consequently, adverse birth outcomes could be hypothesized to be due to placental interface FM tolerance disruption because of cytokine shifts associated with ART initiation causing early initiation of uterine contractions ^20^. Additionally, there are suggestions that adverse birth outcomes could also be associated with ART-induced dysregulation of maternal and infant metabolism. In order to meet the specific needs during pregnancy and infancy, metabolism is tightly regulated, but ART is known to interfere with lipid metabolism ^21^. The emerging field of immunometabolism has shown that alterations in the lipid profile increases susceptibility to viral infections by skewing immune responses towards an inflammatory profile. The complex interplay between pregnancy, HIV/ART, host immunity, adverse birth outcomes and long-term child health is poorly understood as detailed data are sparse and often related to drug combinations which are no longer be in use. Further research is required to examine epidemiologic and immunological associations and inform understandings of underlying biological mechanisms giving rise to adverse birth outcomes.

An increase in PTD rates, and especially of SGA infants, could impact on the long-term growth and development of children, and would have consequences for the health and wellbeing of their families and population more widely. We therefore aimed to improve understanding of maternal immune profiles during pregnancy in the context of ART use during gestation, adverse birth outcomes and long-term child health in Cape Town, South Africa, an area of high HIV prevalence. Our primary focus was to quantify the risk of preterm and SGA deliveries; with our underlying hypotheses that (i) timing of ART use (from before or during pregnancy) is associated with increased risk of PTD, (ii) ART-induced hypertension during pregnancy results in increased risk of SGA and (iii) reconstitution of cellular immune responses during ART in pregnancy results in increases in PTD of appropriate-for-gestational age (AGA) infants. Our secondary focus was to determine long-term (first five years of life) child health outcomes of PTD infants (by weight at gestational age and maternal HIV/ART status). Our hypotheses are that (i) throughout childhood SGA infants are disadvantaged in terms of growth and development compared to preterm AGA infants and (ii) ART use alters maternal and fetal lipid metabolism resulting in susceptibility to infections and alterations in vaccine responses in childhood.

## Cohort Description

### Setting

Between April 2015 and October 2016, we enrolled pregnant women (aged ≥18 years) at their first antenatal care (ANC) visit in a prospective cohort study, at a single large public sector primary care facility (Gugulethu Midwife Obstetric Unit (MOU)) in a low-income high HIV-prevalence sub-district of Cape Town, South Africa.

### Ethical Approval

The study was reviewed and approved by the University of Cape Town Faculty of Health Sciences Human Research Ethics Committee (UCT HREC 739/2014) and the University of Southampton Faculty of Medicine Ethics Committee (12542 PIMS).

### Routine Care Services

As part of routine ANC services, gestational age (GA) was estimated based on date of last menstrual period (LMP) and symphysis-fundal height (SFH). All women without a previous HIV diagnosis underwent HIV testing, with universal ART eligibility. HIV-infected women conceiving while on ART continued their current regimen throughout pregnancy; regimens included NNRTIs such as efavirenz (EFV) or protease inhibitor (PI, predominantly used after failure of first-line therapy). For women initiating ART in pregnancy, a fixed-dose combination of tenofovir (TDF) + emtricitabine (FTC) + efavirenz (EFV) was used throughout.

### Recruitment

Following screening of all women attending their first ANC visit, those ≥18 years were eligible and approached to participate in the study. Women who agreed to participate had their routinely collected LMP-based GA and SFH-based GA reviewed by the counsellor; women estimated to be ≤24 weeks were referred for a research ultrasound scan (US) for formal pregnancy dating by a research sonographer blinded to the midwife assessment. HIV-infected women who were ≤24 weeks’ gestation on US were then recruited into a nested cohort (Group 2); half of these had initiated life-long ART prior to conception and half initiated ART during pregnancy.

All participants provided written informed consent prior to study participation, with re-consenting of mother-infant pairs at the first postpartum visit for paediatric follow-up. Consent for study participation included data abstraction from routine clinical records through the pregnancy and post-partum period.

The overall prospective, observational design includes two ‘nested’ groups of pregnant women:

- **Group 1:** the overall population of pregnant women (≥18 years) seeking ANC services at Gugulethu MOU during a 18-month period; within this group, a subset of women thought to be ≤24 weeks’ gestation based on history or examination (clinical GA) underwent ultrasound scan by a research sonographer for more accurate gestation estimation;
- **Group 2:** all HIV-infected pregnant women seeking ANC who are ≤24 weeks’ gestation at US at their first ANC visit, regardless of current ART use at the first ANC visit (nested within Group 1).

### Participant Baseline Characteristics

A total of 4431 women registered for ANC during the study recruitment period, of whom 4111 (93%) were screened for the study and 3972 (90%) enrolled; all delivered by May 2017 (Figure 1). Main reasons for being screened out were under-age, referrals from Basic Antenatal Clinics, or not interested. Of the enrolled women, 2517 (63.4%) were HIV-uninfected and 1455 (36.6%) HIV-infected (Table 1); 2199 (55.4%) were referred for ultrasound based on their clinical GA, 1327 (60.3%) were HIV-uninfected and 872 (39.7%) HIV-infected.

**Table 1:** Baseline Characteristics of Group 1 women at 1^st^ antenatal care visit (n=3972)

|  | Total<br>N=3972 | HIV-uninfected<br>N=2517 | HIV-infected<br>N=1455 | P value | HIV-infected<br>N= 1455 |  | P value |
| --- | --- | --- | --- | --- | --- | --- | --- |
|  |  |  |  |  | Initiated before<br>pregnancy<br>N=733 | Initiated during<br>pregnancy<br>N=722 |  |
| Age, years |  |  |  | <0.0001 |  |  | <0.0001 |
| <24 | 1273 (32) | 987 (39) | 286 (20) |  | 112 (16) | 174 (24) |  |
| 25-29 | 1108 (28) | 702 (28) | 406 (28) |  | 157 (22) | 249 (34) |  |
| >30 | 1591 (40) | 828 (33) | 763 (52) |  | 453 (63) | 310 (42) |  |
| Median | 28 (23-32) | 26 (22-31) | 30 (25-34) |  | 31 (27–35) | 28 (25-32) |  |
| Height, cm |  |  |  | 0.221 |  |  | 0.562 |
| ≤155 | 1173 (30) | 723 (29) | 450 (31) |  | 220 (30) | 230 (31) |  |
| 156-161 | 1344 (34) | 873 (35) | 471 (32) |  | 245 (34) | 226 (31) |  |
| ≥162 | 1049 (26) | 661 (26) | 388 (27) |  | 190 (27) | 198 (27) |  |
| Missing | 406 (10) | 256 (10) | 147 (10) |  | 67 (9) | 79 (11) |  |
| Median | 158 (154-162) | 158 (154-162) | 158 (154-162) |  | 158 (154-162) | 158 (154-162) |  |
| Body Mass Index, kg/m<sup>2</sup> |  |  |  | 0.666 |  |  | 0.535 |
| Underweight (<18.5) | 26 (0.7) | 15 (0.6) | 11 (0.7) |  | 8 (1) | 3 (0.4) |  |
| Normal (18.5-24.9) | 718 (18) | 457 (18) | 261 (18) |  | 124 (17) | 137 (19) |  |
| Overweight (25.0-29.9) | 1007 (25) | 625 (25) | 381 (26) |  | 202 (28) | 180 (25) |  |
| Obese (>30.0) | 1790 (55) | 1148 (47) | 801 (55) |  | 313 (43) | 329 (55) |  |
| Missing | 431 (11) | 272 (11) | 159 (11) |  | 75 (10) | 84 (11) |  |
| Median | 30 (26-35) | 30 (26-35) | 30 (26-35) |  | 30 (26-35) | 30 (25-35) |  |
| Gravidity |  |  |  | <0.0001 |  |  | 0.002 |
| 1 | 967 (24) | 726 (29) | 241 (17) |  | 100 (14) | 141 (19) |  |
| 2 | 1347 (34) | 849 (34) | 498 (34) |  | 238 (33) | 260 (35) |  |
| ≥3 | 1567 (39) | 887 (35) | 680 (47) |  | 368 (51) | 312 (43) |  |
| Missing | 91 (2) | 55 (2) | 36 (3) |  | 16 (2) | 20 (3) |  |
| Median | 2 (1-3) | 2 (1-3) | 2 (2-3) |  | 3 (2-3) | 2 (2-3) |  |
| Parity |  |  |  | <0.0001 |  |  | 0.011 |
| 0 | 1194 (30) | 865 (34) | 329 (23) |  | 147 (20) | 182 (25) |  |
| 1 | 1458 (37) | 898 (36) | 560 (38) |  | 270 (37) | 290 (40) |  |
| ≥2 | 1228 (31) | 699 (28) | 529 (36) |  | 289 (40) | 240 (33) |  |
| Missing | 92 (2) | 55 (0.1) | 37 (3) |  | 16 (2) | 21 (3) |  |
| Median | 1 (0-2) | 1 (0-2) | 1 (1-2) |  | 1 (1-2) | 1 (0-2) |  |
| Previous Preterm* |  |  |  | 0.013 |  |  | 0.617 |
| Yes | 280 (7) | 159 (6) | 121 (8) |  | 63 (9) | 58 (8) |  |
| Haemoglobin g/dl |  |  |  | <0.0001 |  |  | <0.0001 |
| Normal (≥11.0) | 1446 (36) | 961 (38) | 485 (33) |  | 289 (40) | 196 (27) |  |
| Mild Anaemia (9-10.9) | 1067 (27) | 638 (25) | 429 (29) |  | 177 (25) | 252 (34) |  |
| Moderate Anaemia (7-8.9) | 229 (6) | 109 (4) | 120 (8) |  | 47 (7) | 73 (10) |  |
| Severe Anaemia (<7) | 5 (0.1) | 4 (0.2) | 1 (0.1) |  | 0 | 1 (0.1) |  |
| Missing | 1225 (31) | 805 (32) | 420 (29) |  | 209 (29) | 211 (29) |  |
| Gestational Age Assessment* |  |  |  |  |  |  |  |
| LMP | 3479 (88) | 2219 (88) | 1260 (87) | 0.150 | 642 (89) | 618 (84) | 0.010 |
| Median (weeks) | 17 (12-22) | 17 (12–23) | 16 (11-22) |  | 15 (11-22) | 17 (12-22) |  |
| SFH | 2327 (59) | 1447 (57) | 880 (60) | 0.065 | 403 (56) | 477 (65) | <0.0001 |
| Median (weeks) | 23 (18-28) | 23 (19-28) | 22 (17-27) |  | 22 (16-27) | 23 (18-28) |  |
| US | 2334 (59) | 1411 (56) | 923 (63) | <0.0001 | 433 (60) | 490 (67) | 0.006 |
| Median (weeks) | 16 (12-21) | 16 (12-21) | 15 (11-20) |  | 14 (10-19) | 17 (12-21) |  |

\* LMP – Last menstrual period; SFH – Symphysis fundal height, US – Ultrasound

**Figure 1:**
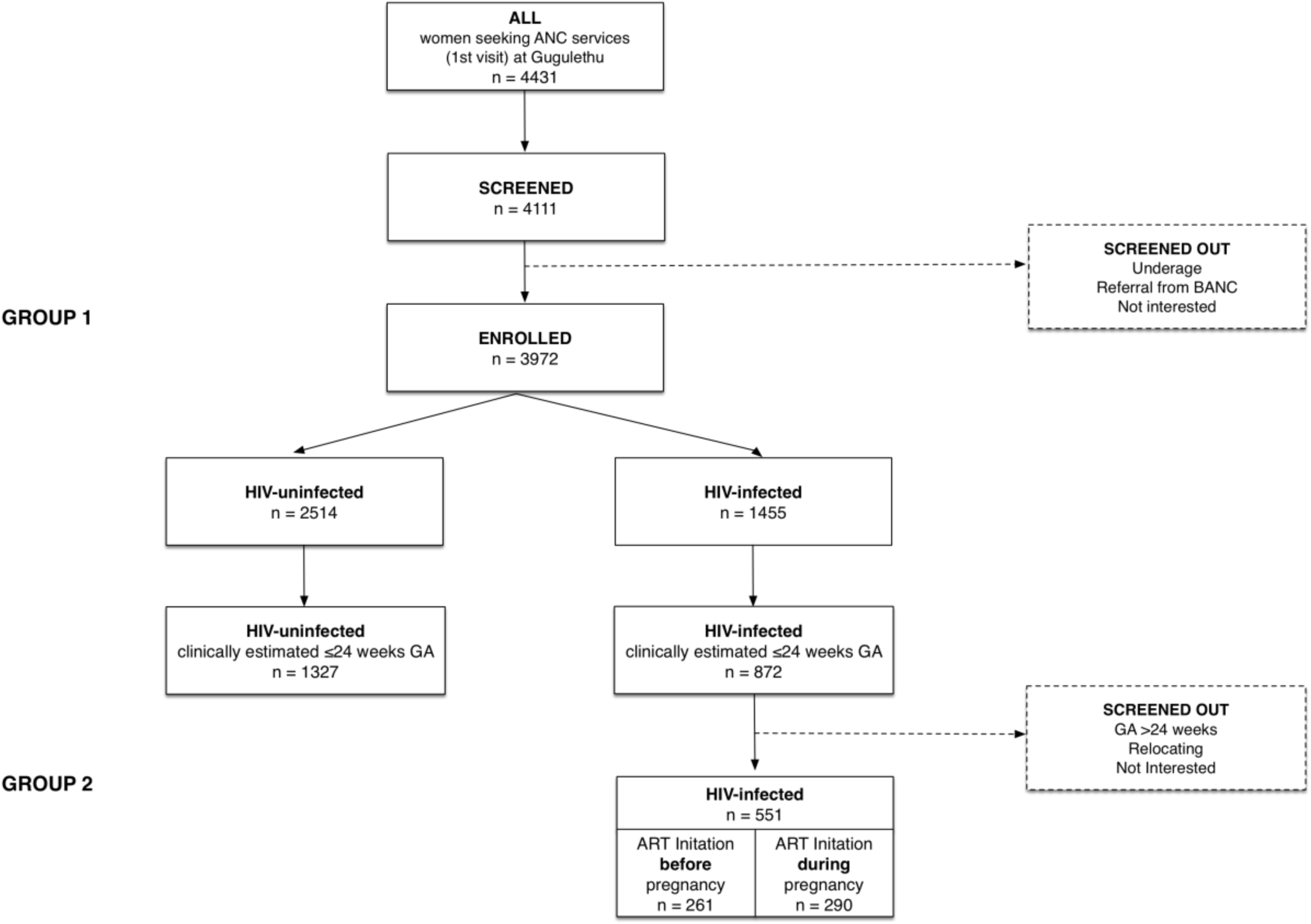
Cohort Profile. BANC – Basic Antenatal Clinic

Median age at enrolment was 28 years (IQR 23-32), with HIV-uninfected women younger than HIV-infected women, and those initiating ART preconception older than HIV-infected women initiating ART during pregnancy. In line with age differences, HIV-infected women were of higher gravidity than HIV-uninfected women, but the difference in parity was small; women who initiated ART pre-pregnancy were of a higher gravidity than those initiating during pregnancy. A quarter (25%) of all women were overweight, while over half (55%) were obese, with little or no difference between groups. Having previously had a preterm delivery was more common among HIV-infected than uninfected women. Mild anaemia was relatively common in all groups, especially in women initiating ART during pregnancy (Table 1). Overall, 3479 (87.6%) women had gestation estimated by LMP, 2327 (58.6%) by SFH and 2334 (58.8%) by ultrasound; with estimated median GA at enrolment visit varying by assessment method (Table 1).

There were 1455 HIV-infected women in Group 1, of whom 718 (49.3%) were ≤24 weeks on ultrasound, and 551 were enrolled into Group 2 (Figure 1). The likelihood of inclusion into Group 2 did not differ by baseline characteristics (Table 2). In comparison to Group 1 participants, Group 2 participants differed in age, gravidity, parity and previous PTD, likely driven by the HIV-uninfected women (Table 3). In multivariable regression allowing for HIV infection, the only difference that persisted between these groups was age (Table 4). When Group 2 women were compared to other HIV-infected women (not enrolled in Group 2), they were slightly older, more likely to have normal haemoglobin levels (≥11g/dl) and lower gestational age (Table 3). In multivariable regression the only difference between these groups that persisted was gestational age, which was a Group 2 inclusion criterion (Table 4).

**Table 2:**
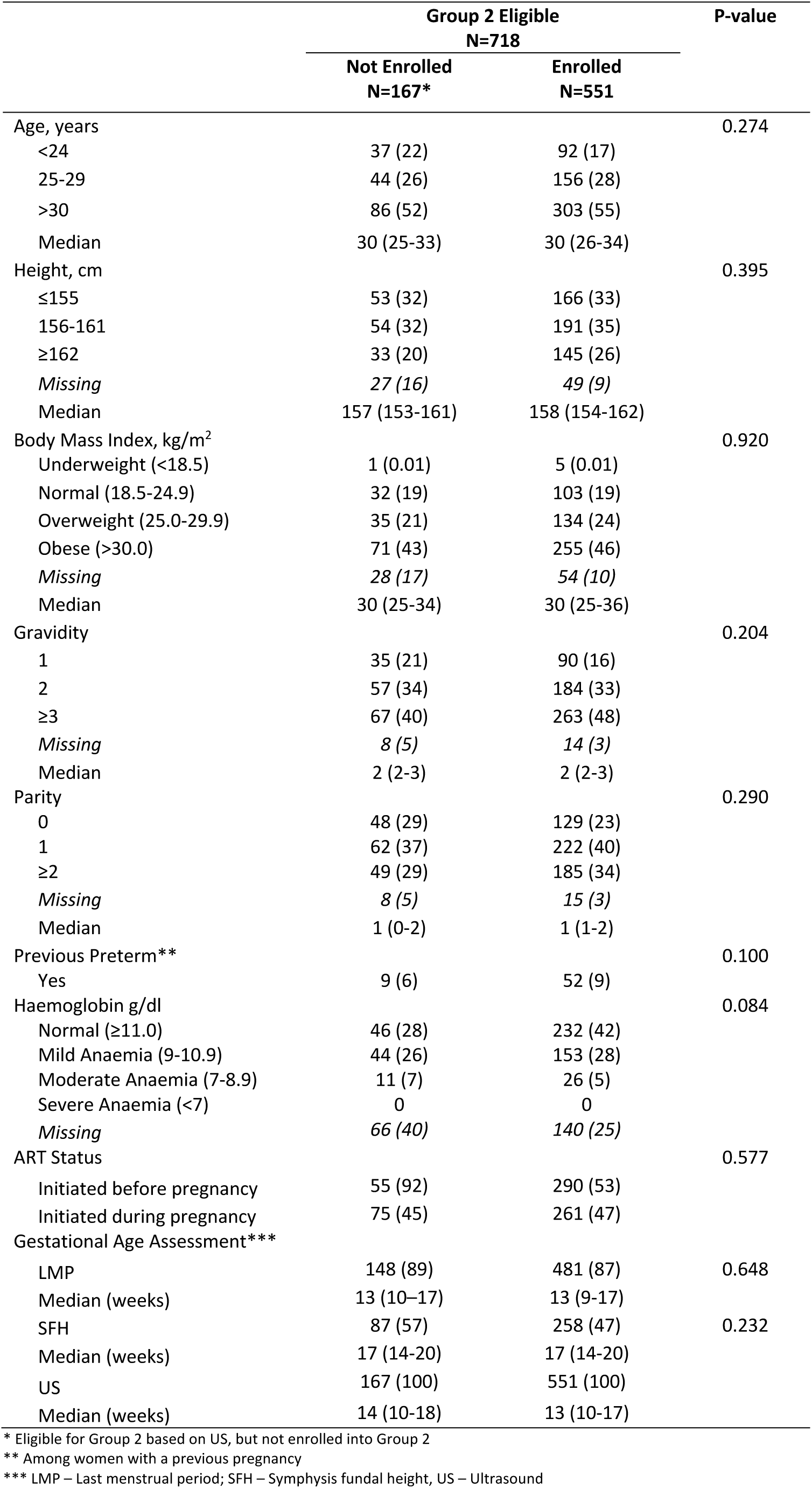
Baseline Characteristics of Group 2 eligible but not enrolled vs Group 2 women at 1^st^ antenatal care visit

**Table 3:** Baseline Characteristics of Group 1 vs Group 2 women at 1^st^ antenatal care visit

|  | Group 1 |  | Group 2 | P-value<br>Grp1 vs Grp2 | P-value<br>HIV+ vs Grp2 |
| --- | --- | --- | --- | --- | --- |
|  | HIV-uninfected<br>N=2517 | HIV-infected*<br>N=904 | HIV-Infected<br>N=551 |  |  |
| Age, years |  |  |  | <0.0001 | 0.078 |
| <24 | 987 (39) | 194 (21) | 92 (17) |  |  |
| 25-29 | 702 (28) | 250 (28) | 156 (28) |  |  |
| >30 | 828 (33) | 460 (51) | 303 (55) |  |  |
| Median | 26 (22-31) | 30 (25-33) | 30 (26-34) |  |  |
| Height, cm |  |  |  | 0.726 | 0.465 |
| ≤155 | 723 (29) | 284 (31) | 166 (30) |  |  |
| 156-161 | 873 (35) | 280 (31) | 191 (35) |  |  |
| ≥162 | 661 (26) | 243 (27) | 145 (26) |  |  |
| Missing | 260 (10) | 97 (11) | 49 (9) |  |  |
| Median | 158 (154-162) | 158 (154-163) | 158 (154-162) |  |  |
| Body Mass Index, kg/m<sup>2</sup> |  |  |  | 0.756 | 0.456 |
| Underweight (<18.5) | 15 (0.6) | 6 (0.7) | 5 (1) |  |  |
| Normal (18.5-24.9) | 457 (18) | 158 (17) | 103 (19) |  |  |
| Overweight (25.0-29.9) | 625 (25) | 248 (27) | 134 (24) |  |  |
| Obese (>30.0) | 1148 (47) | 387 (43) | 255 (46) |  |  |
| Missing | 272 (11) | 105 (12) | 54 (10) |  |  |
| Median | 30 (26-35) | 30 (26-35) | 30 (25-34) |  |  |
| Gravidity |  |  |  | <0.0001 | 0.820 |
| 1 | 726 (29) | 151 (17) | 90 (16) |  |  |
| 2 | 849 (34) | 314 (35) | 184 (33) |  |  |
| ≥3 | 887 (35) | 417 (46) | 263 (48) |  |  |
| Missing | 55 (2) | 22 (2) | 14 (3) |  |  |
| Median | 2 (1-3) | 2 (2-3) | 2 (2-3) |  |  |
| Parity |  |  |  | 0.004 | 0.236 |
| 0 | 865 (34) | 200 (22) | 129 (23) |  |  |
| 1 | 898 (36) | 338 (37) | 222 (40) |  |  |
| ≥2 | 699 (28) | 344 (38) | 185 (34) |  |  |
| Missing | 55 (0.1) | 22 (2) | 15 (3) |  |  |
| Median | 1 (0-2) | 1 (1-2) | 1 (1-2) |  |  |
| Previous Preterm** |  |  |  | <0.0001 | 0.182 |
| Yes | 159 (6) | 69 (8) | 52 (9) |  |  |
| Haemoglobin g/dl |  |  |  | 0.007 | <0.0001 |
| Normal (≥11.0) | 961 (38) | 253 (28) | 232 (42) |  |  |
| Mild Anaemia (9-10.9) | 638 (25) | 276 (31) | 153 (28) |  |  |
| Moderate Anaemia (7-8.9) | 109 (4) | 94 (10) | 26 (5) |  |  |
| Severe Anaemia (<7) | 4 (0.2) | 1 (0.1) | 0 |  |  |
| Missing | 805 (32) | 280 (31) | 140 (25) |  |  |
| Gestational Age Assessment*** |  |  |  |  |  |
| LMP | 2219 (88) | 779 (86) | 481 (87) | 0.823 | 0.542 |
| Median (weeks) | 17 (12-23) | 19 (13-24) | 13 (9-17) |  |  |
| SFH | 1447 (57) | 622 (69) | 258 (47) | <0.0001 | <0.0001 |
| Median (weeks) | 23 (19-28) | 25 (21-29) | 17 (14-20) |  |  |
| US | 1411 (56) | 376 (42) | 551 (100) | <0.0001 | <0.0001 |
| Median (weeks) | 16 (12-21) | 21 (13-25) | 13 (10-17) |  |  |

\* All HIV-infected woman not included in Group 2
\*\* Among women with a previous pregnancy
\*\*\* LMP – Last menstrual period; SFH – Symphysis fundal height, US – ultrasound

**Table 4:** Baseline characteristics associated with inclusion in Group 2 at 1^st^ antenatal care visit

|  | OR<br>Group 1 (Ref) vs Grp 2* |  |  |  | OR<br>Group 1 HIV+ (Ref) vs Group 2** |  |  |  |
| --- | --- | --- | --- | --- | --- | --- | --- | --- |
|  | OR | P-values | aOR (95% CI) | P-values | OR | P-values | aOR (95% CI) | P-values |
| Age, years |  |  |  |  |  |  |  |  |
| <24 | Reference |  | Reference |  | Reference |  | Reference |  |
| 25-29 | 2.10 (1.60-2.76) | <0.0001 | 2.23 (1.64-3.05) | <0.0001 | 1.32 (0.96-1.81) | 0.091 | 1.09 (0.65-1.83) | 0.758 |
| >30 | 3.02 (2.36-3.86) | <0.0001 | 3.37 (2.45-4.63) | <0.0001 | 1.39 (1.04-1.85) | 0.025 | 1.09 (0.66-1.81) | 0.740 |
| Body Mass Index, kg/m<sup>2</sup> |  |  |  |  |  |  |  |  |
| Underweight (<18.5) | Reference |  | Reference |  | Reference |  | Reference |  |
| Normal (18.5-24.9) | 0.70 (0.26-1.91) | 0.489 | 0.75 (0.27-2.06) | 0.573 | 0.78 (0.23-2.63) | 0.691 | 1.02 (0.14-7.37) | 0.988 |
| Overweight (25.0-29.9) | 0.64 (0.24-1.74) | 0.386 | 0.60 (0.22-1.65) | 0.324 | 0.65 (0.19-2.16) | 0.481 | 1.34 (0.19-9.65) | 0.773 |
| Obese (>30.0) | 0.70 (0.26-1.87) | 0.474 | 0.56 (0.20-1.52) | 0.255 | 0.79 (0.24-2.62) | 0.701 | 1.10 (0.15-7.84) | 0.923 |
| Gravidity |  |  |  |  |  |  |  |  |
| 1 | Reference |  | Reference |  | Reference |  | Reference |  |
| 2 | 1.54 (1.18-2.01) | 0.001 | 0.99 (0.73-1.34) | 0.929 | 0.98 (0.72-1.35) | 0.917 | 1.08 (0.64-1.82) | 0.765 |
| ≥3 | 1.97 (1.52-2.53) | <0.0001 | 0.94 (0.68-1.30) | 0.716 | 1.05 (0.78-1.43) | 0.715 | 1.09 (0.64-1.86) | 0.746 |
| Previous Preterm*** |  |  |  |  |  |  |  |  |
| Yes | 1.50 (1.09-2.06) | 0.012 | 1.19 (0.84-1.68) | 0.318 | 1.29 (0.89-1.88) | 0.183 | 1.25 (0.69-2.26) | 0.468 |
| ART Status | - |  | - | - |  |  |  |  |
| Initiated during pregnancy | - |  | - | - | Reference |  | Reference |  |
| Initiated before pregnancy | - |  | - | - | 0.86 (0.70-1.07) | 0.180 | 1.07 (0.76-1.51) | 0.705 |
| Gestational Age | - |  | - | - |  |  |  |  |
| Weeks | - |  | - | - | 0.86 (0.83-0.88) | <0.0001 | 0.84 (0.82-0.87) | <0.0001 |

\* Comparison between all enrolled pregnant women (≥18 years) seeking ANC services at Gugulethu MOU (Group 1) (n=3421) vs HIV-infected pregnant women ≤24 weeks' gestation enrolled into cohort (Group 2) (n=551)
\*\* Comparison between all enrolled HIV-infected pregnant women (≥18 years) seeking ANC services at Gugulethu MOU (Group 1) not enrolled in cohort (n=167) vs HIV-infected pregnant women ≤24 weeks' gestation enrolled into cohort (Group 2) (n=551)
\*\*\* Among women with a previous pregnancy

Of the 551 HIV-infected women in Group 2, 261 (47%) initiated before pregnancy and 290 (53%) during pregnancy (Table 5). Women who initiated ART during pregnancy were on average younger, and of lower gravidity. Overall, three-quarters of Group 2 women were overweight or obese, with little difference by ART status. Of the women who initiated ART during pregnancy, the majority (64%) had tested HIV positive in the index pregnancy; the rest had previously tested positive although were not on ART at conception. In line with local and WHO treatment guidelines, most women (91%) were on a regimen of two NRTIs (TDF +FTC), plus NNRTI EFV. PI usage in this cohort was low, at only 9% of women who initiated pre-pregnancy and 1% of women who initiated during pregnancy. Median CD4 count was 433 cells/µl overall, 527 cells/µl in women who initiated ART before pregnancy and 373 cells/µl in women initiating at the first antenatal visit. There were no differences in smoking or drug usage between these two groups of women, but women who initiated during pregnancy were more likely to have ever consumed alcohol or consumed in the last 30 days (Table 5).

**Table 5:** Baseline Characteristics of Group 2 women at 1^st^ antenatal care visit (n=551)

|  | Total<br>N=551 | HIV-infected |  | P-value |
| --- | --- | --- | --- | --- |
|  |  | Initiation before<br>pregnancy<br>N=261 | Initiation during<br>pregnancy<br>N=290 |  |
| Maternal Characteristics |  |  |  |  |
| Age, years |  |  |  | <0.0001 |
| ≤24 | 92 (17) | 25 (10) | 67 (23) |  |
| 25-29 | 156 (28) | 58 (22) | 98 (34) |  |
| ≥30 | 303 (55) | 178 (68) | 125 (43) |  |
| Median (IQR) | 30 (26-34) | 32 (28-36) | 29 (25-32) |  |
| Education (Finished High School) | 164 (30) | 96 (33) | 69 (26) | 0.088 |
| Employment Status |  |  |  | 0.767 |
| Employed | 238 (46) | 114 (44) | 124 (43) |  |
| Missing | 2 (0.4) | 2 (1) | 0 |  |
| SES |  |  |  | 0.694 |
| Lowest | 175 (32) | 82 (31) | 93 (32) |  |
| Medium | 175 (32) | 88 (34) | 87 (30) |  |
| Highest | 189 (34) | 87 (33) | 102 (35) |  |
| Missing | 12 (2) | 4 (3) | 8 (3) |  |
| Obstetric Characteristics |  |  |  |  |
| Gravidity |  |  |  | <0.0001 |
| 1 | 88 (16) | 29 (11) | 59 (20) |  |
| 2 | 187 (34) | 78 (30) | 109 (38) |  |
| ≥3 | 276 (50) | 154 (59) | 122 (42) |  |
| Median (IQR) | 2 (2-3) | 3 (2-4) | 2 (2-3) |  |
| Parity |  |  |  | 0.061 |
| 0 | 123 (22) | 49 (18) | 74 (26) |  |
| 1 | 220 (40) | 99 (38) | 121 (42) |  |
| ≥2 | 208 (38) | 113 (43) | 95 (33) |  |
| Median (IQR) | 1 (1-2) | 1 (1-2) | 1 (0-2) |  |
| Height, cm |  |  |  | 0.858 |
| ≤155 | 130 (24) | 64 (25) | 66 (23) |  |
| 156-161 | 208 (38) | 96 (37) | 112 (39) |  |
| ≥162 | 208 (38) | 99 (38) | 109 (38) |  |
| Missing | 5 (0.9) | 2 (0.8) | 3 (1) |  |
| Median (IQR) | 160 (156-164) | 159 (155-163) | 160 (156-164) |  |
| Body Mass Index, kg/m² |  |  |  | 0.591 |
| Underweight (<18.5) | 6 (1) | 3 (1) | 3 (1) |  |
| Normal (18.5-24.9) | 110 (20) | 47 (18) | 63 (22) |  |
| Overweight (25.0-29.9) | 148 (27) | 76 (29) | 72 (25) |  |
| Obese (>30.0) | 282 (51) | 133 (51) | 149 (51) |  |
| Missing | 5 (0.9) | 2 (0.8) | 1 (0.3) |  |
| Median (IQR) | 30 (26-35) | 30 (26-34) | 30 (25-35) |  |
| Median Gestation (completed weeks) | 14 (11-18) | 13 (11-17) | 14 (10-18) | 0.054 |
| HIV |  |  |  |  |
| First Tested HIV positive |  |  |  | <0.0001 |
| In this pregnancy | 186 (34) | 0 | 186 (64) |  |
| Before this pregnancy | 365 (66) | 261 (100) | 104 (36) |  |
| ART Use History |  |  |  | <0.0001 |
| Newly Diagnosed | 186 (34) | 0 | 186 (64) |  |
| Known HIV+, No ART | 104 (19) | 0 | 104 (36) |  |
| Known HIV+, On ART | 261 (47) | 261 (100) | 0 |  |
|  |  | Initiation before<br>pregnancy<br>N=261 | Initiation during<br>pregnancy<br>N=290 |  |
| Current ART regimen, self-report |  |  |  | <0.0001 |
| TDF-3TC-EFV | 499 (91) | 220 (84) | 279 (96) |  |
| TDF-3TC-NVP | 4 (1) | 2 (1) | 2 (1) |  |
| Other NNRTI-based regimen | 23 (4) | 16 (6) | 7 (2) |  |
| PI-based regimen | 25 (4) | 23 (9) | 2 (1) |  |
| CD4 cell count, cells/μL* | | | | <0.0001 |
| ≤ 200 | 53 (10) | 13 (5) | 40 (14) |  |
| 201-350 | 111 (20) | 37 (14) | 74 (26) |  |
| 351-500 | 122 (22) | 53 (20) | 69 (24) |  |
| >500 | 194 (34) | 120 (46) | 74 (26) |  |
| Missing | 71 (13) | 38 (15) | 33 (11) |  |
| Median (IQR) | 433 (298-600) | 527 (368-638) | 373 (246-519) |  |
| VL, copies/mL* |  |  |  | 0.015 |
| <400 | 458 (83) | 234 (90) | 224 (77) |  |
| 401-1000 | 14 (3) | 5 (2) | 9 (3) |  |
| >1000 | 64 (12) | 21 (8) | 43 (15) |  |
| Missing | 15 (3) | 1 (0.4) | 14 (5) |  |
| Median (IQR) | 20 (20-67) | 20 (20-20) | 20 (20-100) |  |
| **Substance Use** |  |  |  |  |
| Substance Use, ever |  |  |  |  |
| Alcohol |  |  |  | 0.014 |
| Yes | 357 (65) | 155 (59) | 202 (70) |  |
| No | 189 (34) | 103 (39) | 86 (29) |  |
| Missing | 5 (1) | 3 (1) | 2 (1) |  |
| Smoking |  |  |  | 0.123 |
| Yes | 56 (10) | 21 (8) | 35 (12) |  |
| No | 490 (89) | 237 (91) | 253 (87) |  |
| Missing | 5 (1) | 3 (1) | 2 (1) |  |
| Drugs |  |  |  | 0.146 |
| Yes | 11 (2) | 2 (1) | 9 (3) |  |
| No | 534 (97) | 256 (98) | 278 (96) |  |
| Missing | 6 (1) | 3 (1) | 3 (1) |  |
| Substance Use, last 30 days |  |  |  |  |
| Alcohol |  |  |  | 0.061 |
| Yes | 105 (19) | 41 (16) | 64 (22) |  |
| No | 439 (80) | 216 (83) | 223 (77) |  |
| Missing | 7 (1) | 4 (1) | 3 (1) |  |
| Smoking |  |  |  | 0.101 |
| Yes | 33 (6) | 11 (4) | 22 (7) |  |
| No | 512 (93) | 246 (94) | 266 (92) |  |
| Missing | 6 (1) | 4 (2) | 2 (1) |  |
| Drugs |  |  |  | 0.101 |
| Yes | 3 (1) | 0 | 3 (1) |  |
| No | 542 (98) | 257 (98) | 285 (98) |  |
| Missing | 6 (1) | 4 (2) | 2 (1) |  |

\* CD4 and VL results abstracted from routine records and are the nearest in time to the first ANC visit

### Study Follow-up

At baseline, all women (Group 1) had clinical and medical history, routine first ANC visit physical examination, screening tests and GA assessment data collected via abstraction of the Maternity Case Record (MCR) booklet (Table 5), which is a standardised patient-held maternity record used by all facilities providing maternity services to record clinical data from the antenatal through to postpartum period, including labour. The MCR also serves as a referral letter, thus serving as a link between antenatal and labour care. In addition, the National Health Laboratory Services (NHLS) database was searched for CD4, Viral load results and other laboratory values not recorded in the MCR. Further follow-up for women in Group 1, not eligible for Group 2, was through data abstraction of the MCR following discharge from the postnatal ward (MCR retained at delivery facility). Data was abstracted from follow-up ANC notes, clinical notes during labour and newborn assessments (Table 5).

Women in Group 2 participated in up to eight scheduled study visits, from the start of ANC through to 12 months postpartum. Women on ART from before pregnancy had three antenatal visits at <24 weeks, 28 and 34 weeks; women who initiated ART during pregnancy had an additional study visit two weeks after the ART initiation (which in most women took place on the same day, or close to, the first study visit). Following delivery, women were re-consented for infant participation, and study visits were conducted <7days, 10 weeks, 6 months and 12 months postpartum. At all study visits, data were collected on maternal health (HIV care and ART use, clinical care and inter-current clinical history). Other procedures included physical examinations (anthropometry, blood pressure measurement), phlebotomy (50ml) for storage of plasma, peripheral blood mononuclear cells (PBMC) and RNA; a follow-up ultrasound was conducted at 28 weeks to assess fetal growth (Table 5).

At postpartum study visits, additional data was collected on infant health (including infant feeding and inter-current clinical history) and physical examination of infants was conducted (basic anthropometry). At the 12 month visit, developmental assessment was conducted using the Ages and Stages questionnaire ^22^ – a general developmental screening tool testing five key areas: personal social, gross motor, fine motor, problem solving and communication skills. Infant specimen collection included Dried Blood Spot (DBS) sampling and storage at 10 weeks study visit, and phlebotomy (2ml) for measurement of immunological functioning and antibody responses to routine childhood immunisations (rotavirus and measles) at 12 month study visit. In addition, data on infant health status, including vaccinations, chemoprophylaxis use (including nevirapine and co-trimoxazole) and routine HIV PCR testing, was abstracted from Road-to-Health Booklets - patient-held booklet taken to all clinical and immunisation visits used to monitor infant growth and development until age 5 years (Table 6).

**Table 6:**
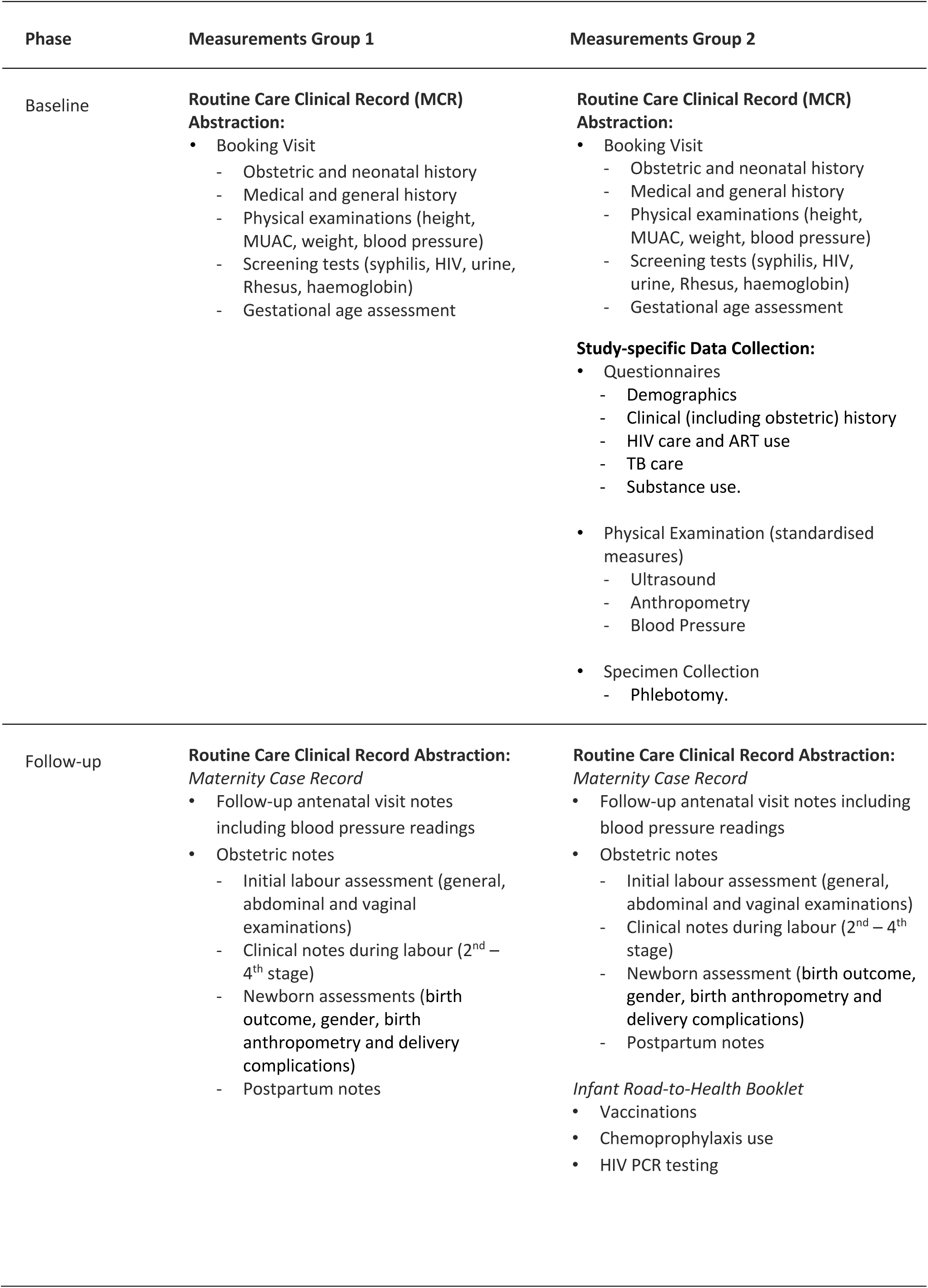
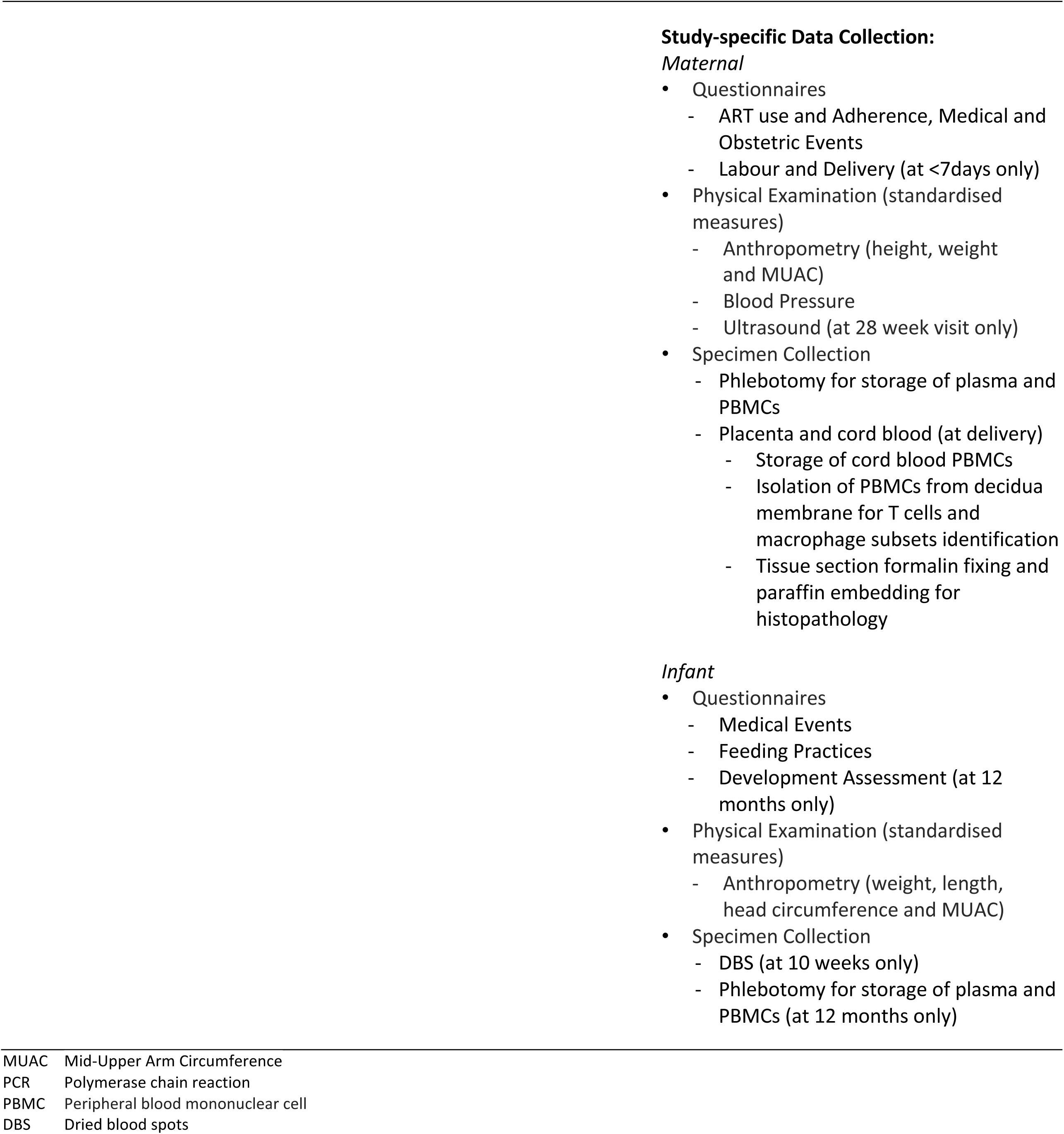
Group 1 and Group 2 Measurements

| Phase | Measurements Group 1 | Measurements Group 2 |
| --- | --- | --- |
| Baseline | **Routine Care Clinical Record (MCR) Abstraction:**<br>• Booking Visit<br>- Obstetric and neonatal history<br>- Medical and general history<br>- Physical examinations (height, MUAC, weight, blood pressure)<br>- Screening tests (syphilis, HIV, urine, Rhesus, haemoglobin)<br>- Gestational age assessment | **Routine Care Clinical Record (MCR) Abstraction:**<br>• Booking Visit<br>- Obstetric and neonatal history<br>- Medical and general history<br>- Physical examinations (height, MUAC, weight, blood pressure)<br>- Screening tests (syphilis, HIV, urine, Rhesus, haemoglobin)<br>- Gestational age assessment<br>**Study-specific Data Collection:**<br>• Questionnaires<br>- Demographics<br>- Clinical (including obstetric) history<br>- HIV care and ART use<br>- TB care<br>- Substance use.<br>• Physical Examination (standardised measures)<br>- Ultrasound<br>- Anthropometry<br>- Blood Pressure<br>• Specimen Collection<br>- Phlebotomy. |
| Follow-up | **Routine Care Clinical Record Abstraction: Maternity Case Record**<br>• Follow-up antenatal visit notes including blood pressure readings<br>• Obstetric notes<br>- Initial labour assessment (general, abdominal and vaginal examinations)<br>- Clinical notes during labour (2<sup>nd</sup> – 4<sup>th</sup> stage)<br>- Newborn assessments (birth outcome, gender, birth anthropometry and delivery complications)<br>- Postpartum notes | **Routine Care Clinical Record Abstraction: Maternity Case Record**<br>• Follow-up antenatal visit notes including blood pressure readings<br>• Obstetric notes<br>- Initial labour assessment (general, abdominal and vaginal examinations)<br>- Clinical notes during labour (2<sup>nd</sup> – 4<sup>th</sup> stage)<br>- Newborn assessment (birth outcome, gender, birth anthropometry and delivery complications)<br>- Postpartum notes<br>*Infant Road-to-Health Booklet*<br>• Vaccinations<br>• Chemoprophylaxis use<br>• HIV PCR testing |

|  |  |
| --- | --- |
| MUAC | Mid-Upper Arm Circumference |
| PCR | Polymerase chain reaction |
| PBMC | Peripheral blood mononuclear cell |
| DBS | Dried blood spots |

Further active follow-up of the women and their infants will occur at school-going age and beyond to chart growth, morbidity and development, as well as changes in family circumstances. It is envisaged that subsequent to this, longer term follow-up will be passive through the use of routinely collected data. The Western Cape Provincial Department of Health’s public-sector patient administration systems all share unique health identifier ^23^; data relating to participants health service contacts, health conditions and health outcomes for specific conditions will be obtained from the Provincial Health Data Centre, which consolidates person-level clinical data across government services ^24^.

### Data Collection

An overview of the main included data collection instruments is presented in Table 6, covering self-reported information on clinical history, ART use and adherence, and medical events; as well as information obtained from routinely collected data.

### Specimen Collection

To investigate the proposed hypothesis that immunological changes resulting from maternal ART exposure are associated with adverse birth outcomes, women enrolled into the follow-up cohort were intensively sampled, with repeated phlebotomy throughout pregnancy and the postpartum period for immunological investigations (Table 7). Using samples from all antenatal plasma, inflammatory markers (C-reactive Protein, Serum Amyloid A and CCL10 (IP-10) in women are being measured. Further, following a nested case-control design in Group 2, investigations compare women delivered preterm (PTD cases) or had small-for-gestational age infants (SGA cases) and those from appropriate controls (term AGA) (matched for GA and ART status). Investigations include longitudinal quantification of plasma cytokine profiles, phenotypic and functional characterisation of regulatory T cells (Tregs), antigen-presenting cells and metabolites associated with mitochondrial functioning and lysophospholipids (Figure 3). The combined studies of these immune parameters will inform understanding of ART use during pregnancy on the areas of the immune system that have been shown to be critically involved in regulating maternal immune tolerance to the fetus, and their associations with onset of labour and preterm delivery.

**Table 7:** Number of available specimens per study visit

| Specimen type | Specimen storage | Visits* |  |  |  |  |  |  |  |  |
| --- | --- | --- | --- | --- | --- | --- | --- | --- | --- | --- |
|  |  | A1 | A1.5 | A2 | A3 | Del | P1 | P2 | P3 | P4 |
| Maternal |  |  |  |  |  |  |  |  |  |  |
| PBMC** | Sodium Heparin | 463 | 227 | 445 | 419 |  | 405 | 412 | 403 | 364 |
|  | EDTA† | 466 | - | - | - |  | 344 | - | - | - |
| Plasma | Sodium Heparin | 483 | 236 | 452 | 424 |  | 407 | 413 | 404 | 366 |
|  | EDTA† | 499 | - | - | - |  | 345 | - | - | - |
|  | PAXGene | 493 | - | - | - |  | - | - | - | - |
| Delivery |  |  |  |  |  |  |  |  |  |  |
| Placenta | Block |  |  |  |  | 229 |  |  |  |  |
|  | OCT†† |  |  |  |  | 190 |  |  |  |  |
|  | RNA Sequencing |  |  |  |  | 176 |  |  |  |  |
|  | RNA later |  |  |  |  | 146 |  |  |  |  |
| Cord Blood |  |  |  |  |  |  |  |  |  |  |
| PBMC** | Sodium Heparin |  |  |  |  | 161 |  |  |  |  |
| Plasma | Sodium Heparin |  |  |  |  | 161 |  |  |  |  |
| Infant |  |  |  |  |  |  |  |  |  |  |
| DBS*** | - |  |  |  |  |  | - | 228 | 67 | 18 |
| PBMC** | Sodium Heparin |  |  |  |  |  | - | - | - | 225 |
| Plasma | Sodium Heparin |  |  |  |  |  | - | - | - | 228 |

\* Study Visits - **A1** Enrolment; **A1.5** ~2 weeks post ART initiation; **A2** ~28 weeks gestation; **A3** ~34 weeks gestation; **P1** ~7 days postpartum, **P2** ~10 weeks postpartum; **P3** ~6 months postpartum; **P4** ~12 months postpartum
\*\* PBMC - Peripheral blood mononuclear cell
\*\*\* DBS - Dried blood spots
† EDTA - Ethylenediaminetetraacetic acid
†† OCT - Optimal cutting temperature

**Figure 2:**
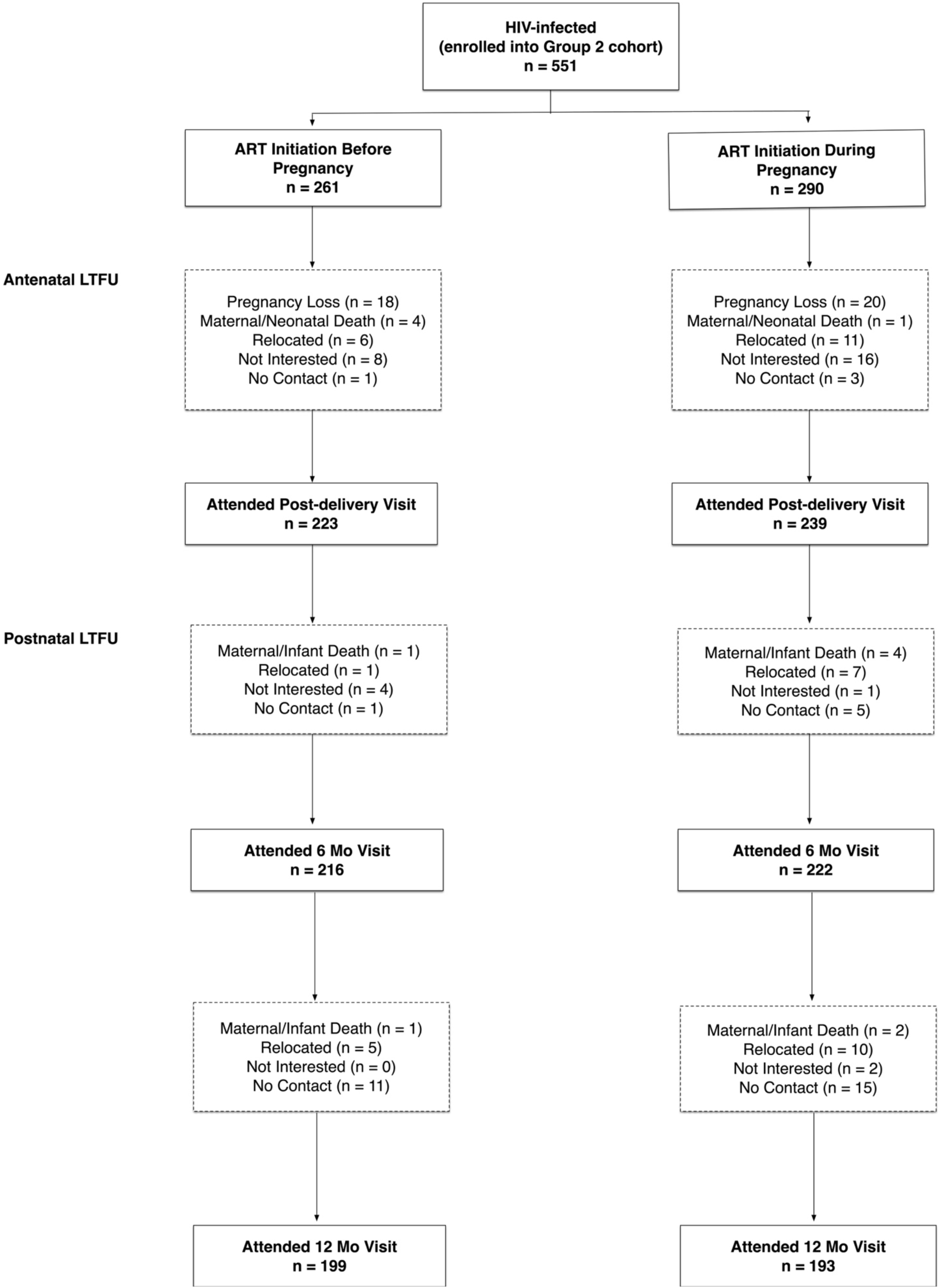
Loss to follow up in Group 2 cohort.

**Figure 3:**
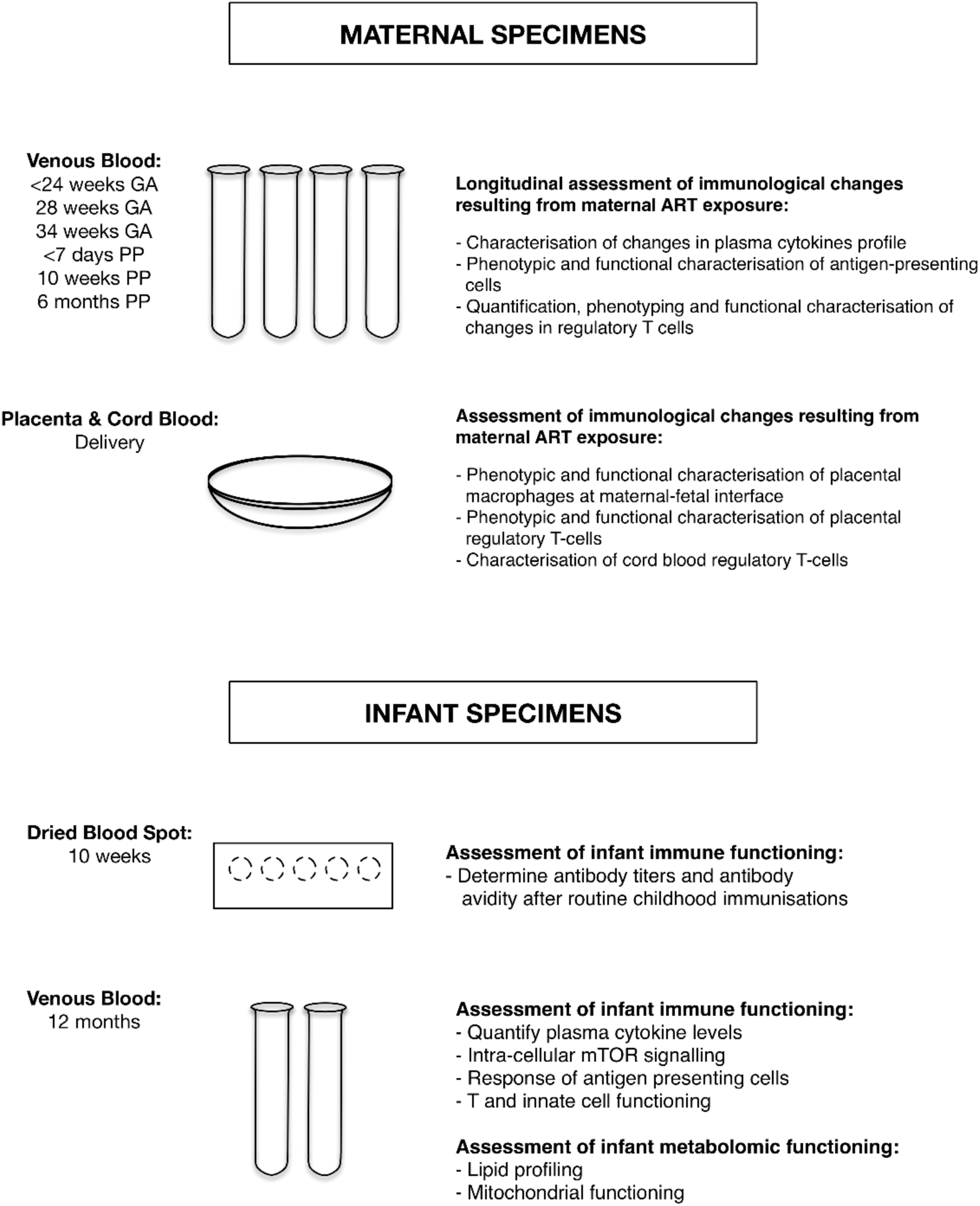
Maternal and Infant Specimen Collection.

At delivery, placentas and cord bloods were collected whenever possible, a scoring algorithm was developed which graded placentas and dictated specimen processing according to membrane completeness and time received in laboratory relative to delivery time (Table 8). Using flow cytometry and tissue immunostaining techniques the following investigations will be conducted: examination of the effect of HIV infection/ART exposure on the phenotypic characteristics and functionality of placental macrophages (Hofbauer cells and decidual macrophages) at the maternal-fetal interface and placental Tregs and their association with adverse birth outcomes. Characterisation of cord blood Tregs and correlation of their frequency, function and phenotype with placental Tregs and birth outcomes (Figure 3).

**Table 8:**
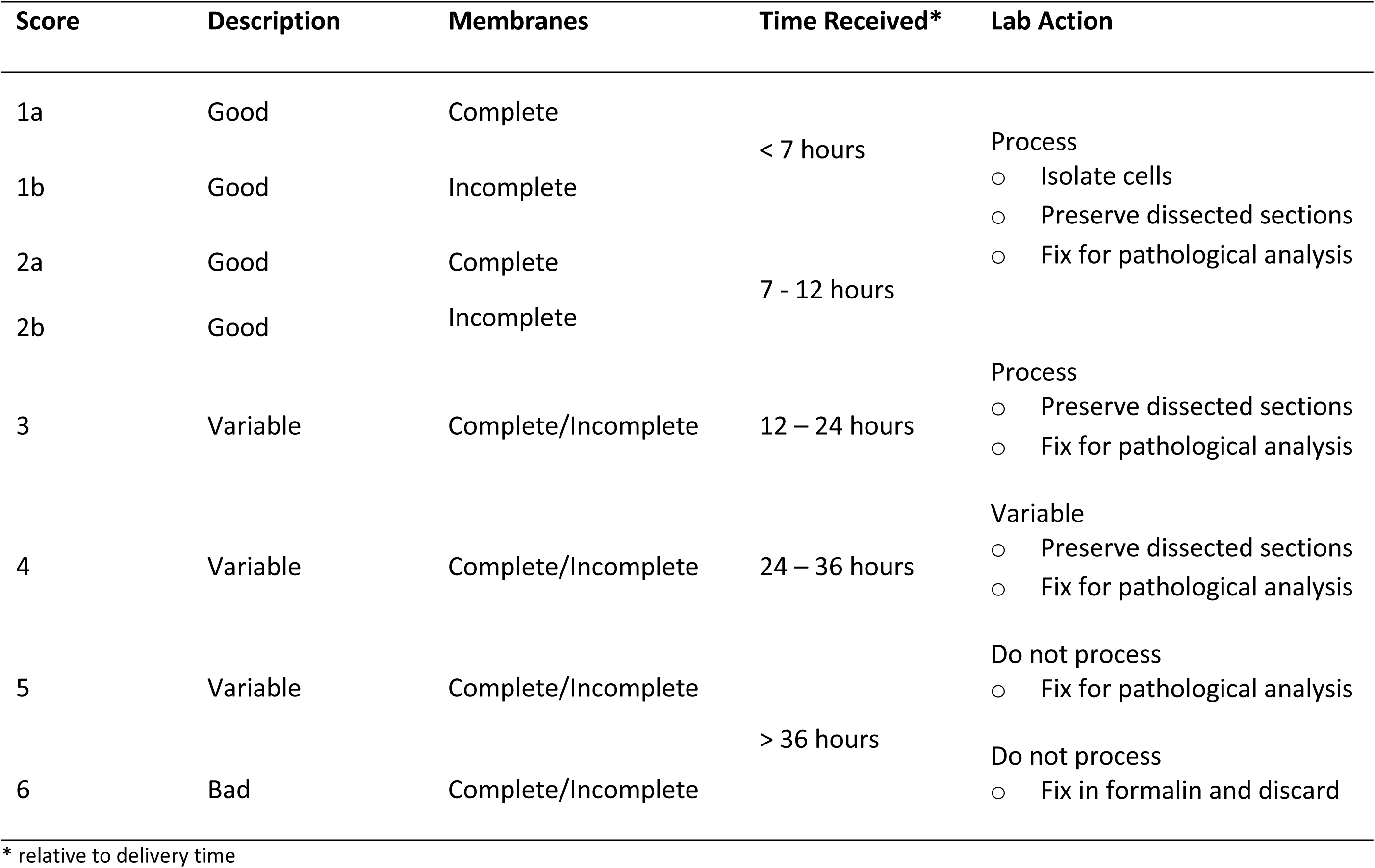
Placenta scoring algorithm for histopathology and laboratory analyses.

### Study Retention

Loss to follow-up was categorised based on the last visit before the woman was lost; in total 158 (29%) women were lost to follow-up (LTFU) (Fig 2). Women lost before the post-delivery study visit (n=88), either experienced pregnancy losses (n=37), were no longer interested in participating (n=24) or relocated out of the study area (n=17). For women LTFU between delivery and the 6 month postpartum visit (n=24), reasons included relocation (n=8), not contactable (n=6), no longer interested in participating (n=5) and maternal/infant death (n=5) (Fig 2). For women LTFU between 6 month and 12 month postpartum (n=46), reasons included relocation (n=15), not contactable (n=26), no longer interested in participating (n=2) and maternal/infant death (n=3).

There were no appreciable differences by ART status in women LTFU before post-delivery visit (RR 0.81, 95% CI 0.55 – 1.19). However, women who initiated ART before pregnancy were less likely to be LTFU between delivery and 6 months postpartum (RR0.44, 95% 0.19 – 1.04) and between 6 month and 12 month postpartum (RR0.58, 95% 0.33 – 1.02). No baseline characteristics were associated with LTFU.

## Findings to date

### Gestational Age Assessment

In the overall cohort, 1787 women with live singleton births were included in the analysis of the association between HIV status and timing of ART initiation and PTD by GA assessment method used (last menstrual period (LMP), measurement of symphysis fundal height (SFH) and ultrasound (US). Using US-GA, PTD risk was associated with maternal HIV infection and ART use, with HIV-infected women, on ART from before or early pregnancy, almost twice as likely to deliver preterm than HIV-uninfected women ^25^. A weaker association was observed when GA assessment was based on SFH; while with LMP-GA the difference by HIV status was minimal. We did not find any appreciable differences in the PTD risk for HIV-infected women by timing of ART initiation across all three assessment methods ^25^. Our findings (in both the overall cohort and in women with all three assessments) suggest that methods of GA assessment explain at least partially the heterogeneity of findings from previous studies on the association between ART use and adverse birth outcomes, suggesting that care should be taken when interpreting results from such studies.

### Obesity

In the overall cohort of HIV-infected and -uninfected women, 2921 women were included in the analysis of the association between maternal body mass index and adverse birth outcomes. In a subset cohort the association between gestational weight gain (GWG) and adverse birth outcomes was examined. Maternal obesity was associated with increased likelihood of having high birthweight and large size for gestational age infants. In the subset cohort, GWG was associated with increased likelihood of spontaneous PTD and high birth weight infants ^26^. Obesity during pregnancy is prevalent in this setting and appears associated with increased risk of adverse birth outcomes in both HIV-infected and -uninfected women.

### Placental Pathology

Preliminary analysis of placental histopathology from a sub-set of women enrolled in the prospective cohort showed significant associations between placental pathology and adverse birth outcomes: presence of focal infarction was associated with increased risk of low birthweight (LBW); the lower the weight of the basal plate weight lead to increased risk of LBW, PTD and SGA; and prolonged meconium exposure was associated with increased risk of SGA. These findings suggest that adverse birth outcomes are driven primarily by placental abnormalities which do not appear to be associated with the ART initiation timing ^27^. Immunofluorescence and immunohistochemistry staining were performed on these wax blocks to identify regulatory T cells along with macrophages. Further analysis is ongoing.

Within the placenta, investigation of the distribution of pro-inflammatory (M1) and anti-inflammatory (M2) placental macrophages at the maternal-fetal interface showed no differences in the tissue density of these macrophages within the decidual membranes and villous tissue according to timing of ART initiation. Data suggest that the Hofbauer Cells (which are fetal macrophages) are not polarized into M1/M2 phenotypes but are rather “intermediate” types ^28^.

## Strengths and limitations

Key strengths of the PIMS study include the recruitment of a large community-based cohort; the observational nature of the study provides good external validity of experiences of a public sector primary care population in pregnancy. A further strength is the use of a research sonographer for the GA assessment in women ≤24 weeks when US is highly reproducible and accurate which is important in studying birth outcomes - compromises in outcome ascertainment methods through the use of imprecise methods can affect the detection of the magnitude of associations.

Additionally, the maternal biological specimens from Group 2 throughout pregnancy is a major strength because it enables immunological, metabolomic and placental investigations which will inform understanding of mechanisms underlying adverse birth outcomes in HIV-infected women. As pregnancy is a state of immunoregulation requiring tolerance of a semi-allogeneic fetus, the assessment of placentas of enrolled women provides a unique opportunity to investigate the link between HIV, ART and adverse birth outcomes. Collection of infant specimens further strengthens the study as it is one of the first studies combining metabolomic and immunologic assessments. This will provide an integrated model of the immune-metabolism association in HIV-exposed infants and the consequences of maternal metabolic dysregulation for the immune responses of the infant. Furthermore, the developmental assessments carried out in the infants provides the opportunity to consider the association between maternal immune function during pregnancy and early childhood immunological and developmental outcomes.

PIMS has valuable sub-designs in addition to the observational study design with the overall cohort stratified by maternal HIV status, ART use, and related risk factors. One sub-design is the cohort study in HIV-infected women, who have data collection through questionnaires, clinical assessments and phlebotomy spanning pregnancy to early infancy. This study design enables quantification of the risk of adverse birth outcomes in the overall cohort, as well as the more detailed Group 2 group also enabling investigation of the consequences of the immune response following ART initiation in pregnancy for onset of labour and preterm delivery. Another sub-design is the nested case–control study which will enable immunological investigations in women who did and did not delivery preterm/SGA infant. The ability to track patients using the Western Cape unique identifier across different health and laboratory services, enables the passive long term follow-up of the Group 2 women and their infants; with available data including patient level data (administrative, demographic and clinical), visit level data (clinical observations and findings), laboratory tests and medication.

A limitation of the study is the lack of an HIV-uninfected or ART-unexposed comparator group for the immunological investigations, which could hinder distinguishing ART exposure from HIV disease. Additionally, some information relating to maternal characteristics was based on self-report and thus subject to potential biases. In order to address this limitation, routinely collected data was also collected to confirm data on birth outcomes.

## Data Availability

The data collected in this study will be available to external investigators interested in collaboration upon submission and approval of a data analysis plan. The samples are being used by the named investigators, but remaining samples can be made available to external users. Requests for data and available samples within PIMS, or to submit a request for additional data collection, should be submitted to and will be reviewed by the study steering committee.

## Further Details

Research reported in this publication was supported by the Eunice Kennedy Shriver National Institute of Child Health & Human Development of the National Institutes of Health under Award Number R01HD080385. The content is solely the responsibility of the authors and does not necessarily represent the official views of the National Institutes of Health.

## Acknowledgements

The authors would like to thank all PIMS participants for their valuable contribution to this study.

## Notes

### Competing Interest Statement

The authors have declared no competing interest.

### Funding Statement

The research reported in this publication was supported by the Eunice Kennedy Shriver National Institute of Child Health & Human Development of the National Institutes of Health under Award Number R01HD080385. The content is solely the responsibility of the authors and does not necessarily represent the official views of the National Institutes of Health.

